# Patterns of antidepressant prescribing in and around pregnancy: a descriptive analysis in the UK Clinical Practice Research Datalink

**DOI:** 10.1101/2024.08.08.24311553

**Authors:** Florence Z. Martin, Gemma C. Sharp, Kayleigh E. Easey, Paul Madley-Dowd, Liza Bowen, Victoria Nimmo-Smith, Aws Sadik, Jonathan L. Richardson, Dheeraj Rai, Harriet Forbes

## Abstract

**Objective:** To describe the prevalence and patterns of antidepressant prescribing in and around pregnancy.

**Design:** Drug utilisation study.

**Setting:** Primary care in the United Kingdom (UK).

**Population:** Women with a pregnancy between 1996 and 2018 in the UK Clinical Practice Research Datalink (CPRD) GOLD Pregnancy Register.

**Methods:** Using primary care prescription records, we identified individuals who had been prescribed antidepressants in and around pregnancy and described changing prevalence of prescribing during pregnancy over time. We defined ‘prevalent’ or ‘incident’ antidepressant use, where ‘prevalent’ users were prescribed antidepressants both before and during pregnancy, and ‘incident’ users were newly prescribed antidepressants during pregnancy, then compared patterns of prescribing between these two groups. We also investigated characteristics associated with antidepressant discontinuation anytime during pregnancy and post-pregnancy prescribing.

**Main outcome measures:** Antidepressant prescribing during pregnancy.

**Results:** A total of 1,033,783 pregnancies were identified: 79,144 (7.7%) were prescribed antidepressants during pregnancy and 15,733 of these (19.9%) were ‘incident’ users. Antidepressant prescribing during pregnancy increased from 3.2% in 1996 to 13.4% in 2018. Most women, both ‘prevalent’ and ‘incident’ users, discontinued antidepressants anytime during pregnancy (54.8% and 59.9%, respectively). The majority of those who discontinued during pregnancy resumed in the 12 months after pregnancy (53.0%). Younger age, previous stillbirth, and higher deprivation were associated with more frequent discontinuation anytime during pregnancy.

**Conclusions:** Antidepressant use during pregnancy appears to be increasing in the UK. Most women discontinued antidepressants at some point before the end of pregnancy, but post-pregnancy resumption of antidepressants was common.

**Funding:** Wellcome Trust 218495/Z/19/Z.

## 1 Introduction

Antidepressants are widely prescribed medications^1, 2^ and are used for a range of indications, predominantly depression and anxiety.^3, 4^ Pregnancy is not a contraindication for antidepressants; however, the UK’s National Institute for Health and Care Excellence (NICE) lays out a series of recommendations for depression management before, during, and after pregnancy.^5–8^ The guidance refers to regimen changes such as discontinuation, dose tapering, and product switching if the risk of maternal condition destabilisation is lower than the potential risk to the fetus, assessed on an individual basis.^5, 6^ “Risk to the fetus” refers to both the uncertain effects of the medication *in utero*^9–11^ and potential consequences of unmanaged maternal illness on the fetus, via physiological imbalances or characteristic differences, such as smoking and poor diet.^12–14^

Antidepressant use is increasing globally among women of a childbearing age outside of pregnancy.^15–18^ During pregnancy, previous data from the UK (excluding Wales) suggested that 3.7% of women who had a delivery (either a live- or stillbirth) between 2004 and 2010 were exposed to selective serotonin reuptake inhibitors (SSRIs) during pregnancy, dropping from 8.8% in the year before pregnancy.^19^ This drop may reflect the clinical guidance, where discontinuation has been recommended,^5^ or reflect stigmatisation of antidepressant use during pregnancy and limited evidence for their safety.^20^ Similar patterns of discontinuation have been found in a previous study of antidepressant use during pregnancy.^21^ As guidelines are updated based on emerging evidence, so do prescribing patterns, and it is important to monitor them for research and clinical purposes.

The Clinical Practice Research Datalink (CPRD) GOLD is a repository of primary care data from the UK that pulls data from practices that use the Vision software to collect patient data.^22^ Previous antidepressant utilisation during pregnancy studies have used the CPRD GOLD Mother-Baby Link, which includes all mother-baby pairs where both the mother and baby were registered with a CPRD GOLD practice.^19, 21^ Here instead, we used CPRD GOLD’s Pregnancy Register, capturing all pregnancy episodes in the CPRD GOLD population, regardless of delivery type or child registration with a CPRD GOLD practice.^23^ We show the trend of antidepressant prescribing during pregnancy between 1996 and 2018, patterns of antidepressant prescribing in and around pregnancy, and characteristics associated with discontinuation during pregnancy.

## 2 Methods

The present drug utilisation study leveraged UK electronic health records; all scripts can be accessed via https://github.com/flozoemartin/Patterns.

### 2.1 Data sources

CPRD GOLD consists of primary care data from consenting general practices (GPs) that use the Vision software.^22^ It covers around 7% of the UK population and is broadly representative by age, sex, and ethnicity.^22^ CPRD GOLD contains information on prescriptions using British National Formulary (BNF) codes and diagnoses using Read codes. The CPRD GOLD Pregnancy Register contains algorithmically derived information on all pregnancy episodes in the CPRD GOLD population.^23^ CPRD GOLD is linked to external data sources, including Hospital Episode Statistics (HES) using International Classification of Diseases Tenth Revision (ICD-10) codes, Office for National Statistics (ONS), and Index of Multiple Deprivation (IMD)^22^ (Methods S1.1).

All data in CPRD GOLD are pseudonymised which precludes the need for patient consent and details of CPRD’s safeguarding processes can be found at https://cprd.com/safeguarding-patient-data. Patient and public engagement was not performed as part of this study.

### 2.2 Population

Using the CPRD GOLD Pregnancy Register, we included patients with an estimated pregnancy start date between January 1^st^, 1996, and December 31^st^, 2018, that ended in either a loss or delivery. Eligibility included registration with an ‘up-to-standard’ (UTS) practice for at least 12 months prior to pregnancy start until the end of pregnancy,^22^ allowing sufficient time for collection of information prior to pregnancy. The unit of measurement here is a pregnancy; multiple pregnancies were included and considered once and individuals who had more than one eligible pregnancy were included for each pregnancy. Unknown outcomes and conflicting pregnancies were resolved where possible as per Campbell *et al.*’s approach^24^ (Methods S1.2); unresolved pregnancies were dropped.

### 2.3 Antidepressant prescribing

Antidepressants were defined using validated codelists and divided into SSRIs, serotonin-noradrenaline reuptake inhibitors (SNRIs), tricyclic antidepressants (TCAs), and ‘other’ antidepressants (Table S1). We identified prescriptions made in primary care in the 12 months before, during, and in the 12 months after pregnancy.

Prescription length was calculated by dividing the quantity of tablets prescribed by the number of tablets to be taken each day to estimate the prescription end date. We used hot-decking imputation^25^ where these were missing. We calculated daily dose in milligrams by multiplying the number of tablets prescribed per day by the number of milligrams delivered per dose. Daily dose in milligrams for each medication was then standardised to low, medium, or high based on dose distributions (Methods S1.3). Individuals with prescriptions of different products that overlapped by more than four weeks were defined as being on a ‘multi-drug regimen’.

Exposure to antidepressants was defined as a prescription overlapping with the period of interest: the 12 months prior to pregnancy, during pregnancy (each week of gestation), and the 12 months post-pregnancy.

Pre-pregnancy discontinuation was defined by prescription for antidepressants in the 12 months before pregnancy but not during pregnancy (Figure S1).

Discontinuation during pregnancy was defined as antidepressant prescribing ending more than 2 weeks prior to the end of pregnancy. Discontinuation by trimester is described below (Methods S1.4).

Among those who continued a single-drug regimen, antidepressant switching and dose changes were characterised; among those that continued a multi-drug regimen, product adding, product dropping, and dose changes were characterised (Figure S2, Methods S1.4).

The above patterns were explored among all individuals prescribed antidepressants, as well as ‘prevalent’ and ‘incident’ users. ‘Prevalent’ users had at least one prescription in the 3 months prior to pregnancy and during pregnancy. ‘Incident’ users had no antidepressant prescriptions in the 3 months before pregnancy and at least one antidepressant prescription any time during pregnancy (Figure S1).

Antidepressant prescribing after pregnancy was defined as those with at least one prescription in the 12 months post-pregnancy. Post-pregnancy prescribing was stratified by new users and discontinuers from before and during pregnancy.

### 2.4 Indication

Using the BNF and European Medicines Agency, we identified indications for antidepressants in the UK as of 2023. Read and ICD-10 codes were applied to primary and secondary care data (where available) to identify depression, anxiety, other mood disorders with a depressive element, eating disorders, pain, diabetic neuropathy, stress (urinary) incontinence, migraine prophylaxis, tension-type headache, and narcolepsy with cataplexy, any time prior to or during pregnancy (Methods S1.5).

### 2.5 Characteristics

Information on maternal characteristics was abstracted on all eligible pregnancies. Demographics such as age (at the start of pregnancy), body mass index (BMI, around the start of pregnancy), ethnicity, socioeconomic position (SEP) (proxied using practice-level IMD quintile),^26^ gravidity and parity (at the start of pregnancy), primary care consultations (in the 12 months before pregnancy), prescriptions of other medications (in the 12 months before pregnancy), and other diagnoses (ever before the start of pregnancy), were captured from relevant data sources (Table S2).

### 2.6 Statistical analysis

#### 2.6.1 TRENDS

We calculated the proportion of pregnancies in each year prescribed antidepressants during pregnancy and restricted to pregnancies ending in live births in sensitivity analysis. We showed antidepressant prescribing during pregnancy by UK region across the study period.

#### 2.6.2 Patterns

Among eligible pregnancies, we calculated the proportion of individuals who were prescribed antidepressants prior to pregnancy and of these, the proportion who discontinued prior to pregnancy.

Of those prescribed antidepressants during pregnancy, we described discontinuation and continuation of a single- and multi-drug regimen (i.e., switching and dose changes), repeating analyses stratified by ‘prevalent’ and ‘incident’ users. We explored trimester of discontinuation by restricting to those with at least 29 completed weeks’ gestation. We stratified the primary patterns analysis by delivery type, either deliveries or losses, and restricted to those with linked secondary care data.

Of those who were prescribed antidepressants after pregnancy, we calculated the proportion who were new users and who were resuming having discontinued prior to or during pregnancy. To explore post-pregnancy compared to postnatal prescribing, we stratified this analysis by delivery type in sensitivity analysis. We also restricted the sample to those with at least 12 months post-pregnancy follow-up as we didn’t impose a post-pregnancy follow-up eligibility criterion.

#### 2.6.3 Indications

We characterised timing of depression and anxiety and calculated the proportion of those prescribed antidepressants during pregnancy that had diagnoses of each identified indication.

#### 2.6.4 Predictors of discontinuation

Logistic regression, minimally adjusted for pregnancy start year, was used to understand the relationship between patient characteristics and antidepressant discontinuation anytime during pregnancy. Each logistic regression model was a complete records analysis (CRA), so patients were dropped in the event of missing data. We ran a sensitivity analysis investigating the association between record missingness and discontinuation during pregnancy to assess the potential bias introduced in a CRA.^27^

All analyses were performed in Stata 17 and R 4.3.1. This study was approved by CPRD’s Independent Scientific Advisory Committee (ISAC) in 2021 [ISAC number: 21_000362].

## 3 Results

### 3.1 Study population

Of the pregnancies in the CPRD Pregnancy Register (September 2021), 1,033,783 were eligible (Figure S3). Reasons for ineligibility included the pregnancy occurring outside the study window (*n*=3,276,401) or women having insufficient follow-up with a UTS practice (*n*=2,271,333) (Figure S3). Most pregnancies in the sample ended in live birth (71.1%), with 12.3% ending in miscarriage and 13.5% ending in termination, among other outcomes (Table S3).

Among eligible pregnancies over the entire study period, 142,817 (13.8%) were prescribed antidepressants in the 12 months prior to pregnancy and 79,144 (7.7%) were prescribed antidepressants anytime during pregnancy.

Women prescribed antidepressants during pregnancy were more likely to smoke (43.4% versus 28.7%) and live in the most deprived IMD quintile (30.6% versus 26.9%) than non-prescribed. Other mental health-related prescriptions were more commonly prescribed to women who were prescribed antidepressants during pregnancy (e.g., mood stabilisers were prescribed for 6.7% of prescribed versus 0.7% of non-prescribed). High-dose folic acid and anti-emetics were more widely prescribed during pregnancy to those also prescribed antidepressants than those who were not (Table 1).

**Table 1.** Characteristics table.

| Maternal characteristics | Total | Prescribed antidepressants during pregnancy | Not prescribed antidepressants during pregnancy |
| --- | --- | --- | --- |
| **Total** | 1,033,783 (100.0) | 79,144 (100.0) | 954,639 (100.0) |
| **Maternal age at start of pregnancy** |  |  |  |
| 18 or younger | 38,836 (3.8) | 1,117 (1.4) | 37,719 (4.0) |
| 18–24 | 234,583 (22.7) | 19,196 (24.3) | 215,387 (22.6) |
| 25–29 | 265,993 (25.7) | 20,632 (26.1) | 245,361 (25.7) |
| 30–34 | 287,482 (27.8) | 20,545 (26.0) | 266,937 (28.0) |
| 35 or older | 206,889 (20.0) | 17,654 (22.3) | 189,235 (19.8) |
| **Practice Index of Multiple Deprivation (IMD)** |  |  |  |
| 1<sup>st</sup> quintile (least deprived) | 163,727 (15.8) | 10,316 (13.0) | 153,411 (16.1) |
| 2<sup>nd</sup> quintile | 167,765 (16.2) | 11,811 (14.9) | 155,954 (16.3) |
| 3<sup>rd</sup> quintile | 189,474 (18.3) | 14,390 (18.2) | 175,084 (18.3) |
| 4<sup>th</sup> quintile | 231,787 (22.4) | 18,433 (23.3) | 213,354 (22.3) |
| 5<sup>th</sup> quintile (most deprived) | 281,030 (27.2) | 24,194 (30.6) | 256,836 (26.9) |
| **Maternal ethnicity** |  |  |  |
| White | 639,193 (61.8) | 51,322 (64.8) | 587,871 (61.6) |
| South Asian | 31,837 (3.1) | 959 (1.2) | 30,878 (3.2) |
| Black | 16,920 (1.6) | 513 (0.6) | 16,407 (1.7) |
| Other | 11,235 (1.1) | 357 (0.5) | 10,878 (1.1) |
| Mixed | 6,657 (0.6) | 429 (0.5) | 6,228 (0.7) |
| Missing | 327,941 (31.7) | 25,564 (32.3) | 302,377 (31.7) |
| **Maternal body mass index (kg/m<sup>2</sup>) at start of or around pregnancy** |  |  |  |
| Underweight (<18.5) | 33,926 (3.3) | 2,890 (3.7) | 31,036 (3.3) |
| Healthy weight (18.5–24.9) | 470,954 (45.6) | 30,986 (39.2) | 439,968 (46.1) |
| Overweight (25.0–29.9) | 241,326 (23.3) | 18,783 (23.7) | 222,543 (23.3) |
| Obese (30.0) | 181,912 (17.6) | 19,949 (25.2) | 161,963 (17.0) |
| Missing | 105,665 (10.2) | 6,536 (8.3) | 99,129 (10.4) |
| **Maternal smoking status in and around pregnancy** |  |  |  |
| Non-smoker | 419,921 (40.6) | 21,634 (27.3) | 398,287 (41.7) |
| Ex-smoker | 251,655 (24.3) | 20,905 (26.4) | 230,750 (24.2) |
| Current smoker | 308,267 (29.8) | 34,362 (43.4) | 273,905 (28.7) |
| Missing | 53,940 (5.2) | 2,243 (2.8) | 51,697 (5.4) |
| **Maternal alcohol intake in and around pregnancy** |  |  |  |
| Non-drinker | 133,669 (12.9) | 9,915 (12.5) | 123,754 (13.0) |
| Current drinker | 594,192 (57.5) | 45,315 (57.3) | 548,877 (57.5) |
| Ex-drinker | 60,462 (5.8) | 6,618 (8.4) | 53,844 (5.6) |
| Missing | 245,460 (23.7) | 17,296 (21.9) | 228,164 (23.9) |
| **Maternal illicit drug use prior to or during pregnancy** |  |  |  |
| Yes | 2,378 (0.2) | 914 (1.2) | 1,464 (0.2) |
| **Maternal history of pregnancy loss at the start of pregnancy** |  |  |  |
| Miscarriage | 162,414 (15.7) | 15,405 (19.5) | 147,009 (15.4) |
| Stillbirth | 6,345 (0.6) | 722 (0.9) | 5,623 (0.6) |
| Termination | 174,264 (16.9) | 19,360 (24.5) | 154,904 (16.2) |
| Other losses | 15,969 (1.5) | 1,617 (2.0) | 14,352 (1.5) |
| **Maternal parity at the start of pregnancy** |  |  |  |
| 0 | 489,830 (47.4) | 29,080 (36.7) | 460,750 (48.3) |
| 1 | 348,858 (33.7) | 26,667 (33.7) | 322,191 (33.8) |
| 2 | 132,186 (12.8) | 14,754 (18.6) | 117,432 (12.3) |
| 3 or more | 58,465 (5.7) | 8,324 (10.5) | 50,141 (5.3) |
| **GP consultations in the 12 months before pregnancy** |  |  |  |
| 0 | 116,218 (11.2) | 5,177 (6.5) | 111,041 (11.6) |
| 1–3 | 266,266 (25.8) | 5,291 (6.7) | 260,975 (27.3) |
| 4–9 | 426,917 (41.3) | 26,633 (33.7) | 400,284 (41.9) |
| 10 or more | 224,382 (21.7) | 42,043 (53.1) | 182,339 (19.1) |
| **Mental health problems ever before the end of pregnancy** |  |  |  |
| Depression | 263,063 (25.4) | 63,795 (80.6) | 199,268 (20.9) |
| Anxiety | 164,115 (15.9) | 38,266 (48.3) | 125,849 (13.2) |
| Bipolar (depressive episodes) | 2,498 (0.2) | 888 (1.1) | 1,610 (0.2) |
| Schizophrenia | 2,405 (0.2) | 883 (1.1) | 1,522 (0.2) |
| Eating disorders<sup>3</sup> | 19,635 (1.9) | 4,142 (5.2) | 15,493 (1.6) |
| **Maternal neurodevelopmental disorder ever before the end of pregnancy** |  |  |  |
| Autism spectrum disorder (ASD) | 312 (0.0) | 74 (0.1) | 238 (0.0) |
| Attention deficit hyperactivity disorder (ADHD) | 1,822 (0.2) | 361 (0.5) | 1,461 (0.2) |
| Intellectual disability (ID) | 1,135 (0.1) | 206 (0.3) | 929 (0.1) |
| **Possible somatic indications for antidepressants ever before the end of pregnancy** |  |  |  |
| Neuropathic pain/fibromyalgia | 48,086 (4.7) | 7,263 (9.2) | 40,823 (4.3) |
| Diabetic neuropathy | 93 (0.0) | 28 (0.0) | 65 (0.0) |
| Stress (urinary) incontinence | 13,661 (1.3) | 2,213 (2.8) | 11,448 (1.2) |
| Migraine prophylaxis | 3,552 (0.3) | 638 (0.8) | 2,914 (0.3) |
| Chronic tension-type headache | 418 (0.0) | 88 (0.1) | 330 (0.0) |
| **Other mental health-related prescriptions during pregnancy** |  |  |  |
| Antipsychotics | 579 (0.1) | 392 (0.5) | 187 (0.0) |
| Mood stabilisers | 11,618 (1.1) | 5,265 (6.7) | 6,353 (0.7) |
| Benzodiazepines | 4,989 (0.5) | 2,968 (3.8) | 2,021 (0.2) |
| Z-drugs | 7,768 (0.8) | 2,438 (3.1) | 5,330 (0.6) |
| **Other prescriptions during pregnancy** |  |  |  |
| Other drugs not recommended by obstetricians<sup>4</sup> | 50,478 (4.9) | 9,141 (11.5) | 41,337 (4.3) |
| Folic acid as a multivitamin | 212,664 (20.6) | 19,104 (24.1) | 193,560 (20.3) |
| High dose folic acid (5mg) | 52,724 (5.1) | 6,776 (8.6) | 45,948 (4.8) |
| Antiemetics | 82,816 (8.0) | 13,202 (16.7) | 69,614 (7.3) |

<sup>1</sup> A prescription made during or overlapping with pregnancy
<sup>2</sup> No prescription made during or overlapping with pregnancy
<sup>3</sup> Anorexia nervosa, bulimia, and other disordered eating codes (codelist in the supplementary material)
<sup>4</sup> “Classical teratogens” like isotretinoin and ACE inhibitors (codelist in the supplementary material)

### 3.2 Trends over time

Antidepressant prescribing during pregnancy increased from 3.2% in 1996 to 13.4% in 2018. Exclusive treatment with SSRIs has dominated antidepressant prescribing during pregnancy (Figure 1). We observed a similar increase when restricting to live births (2.6% in 1996 to 12.6% in 2018) (Figure S4).

**Figure 1.**
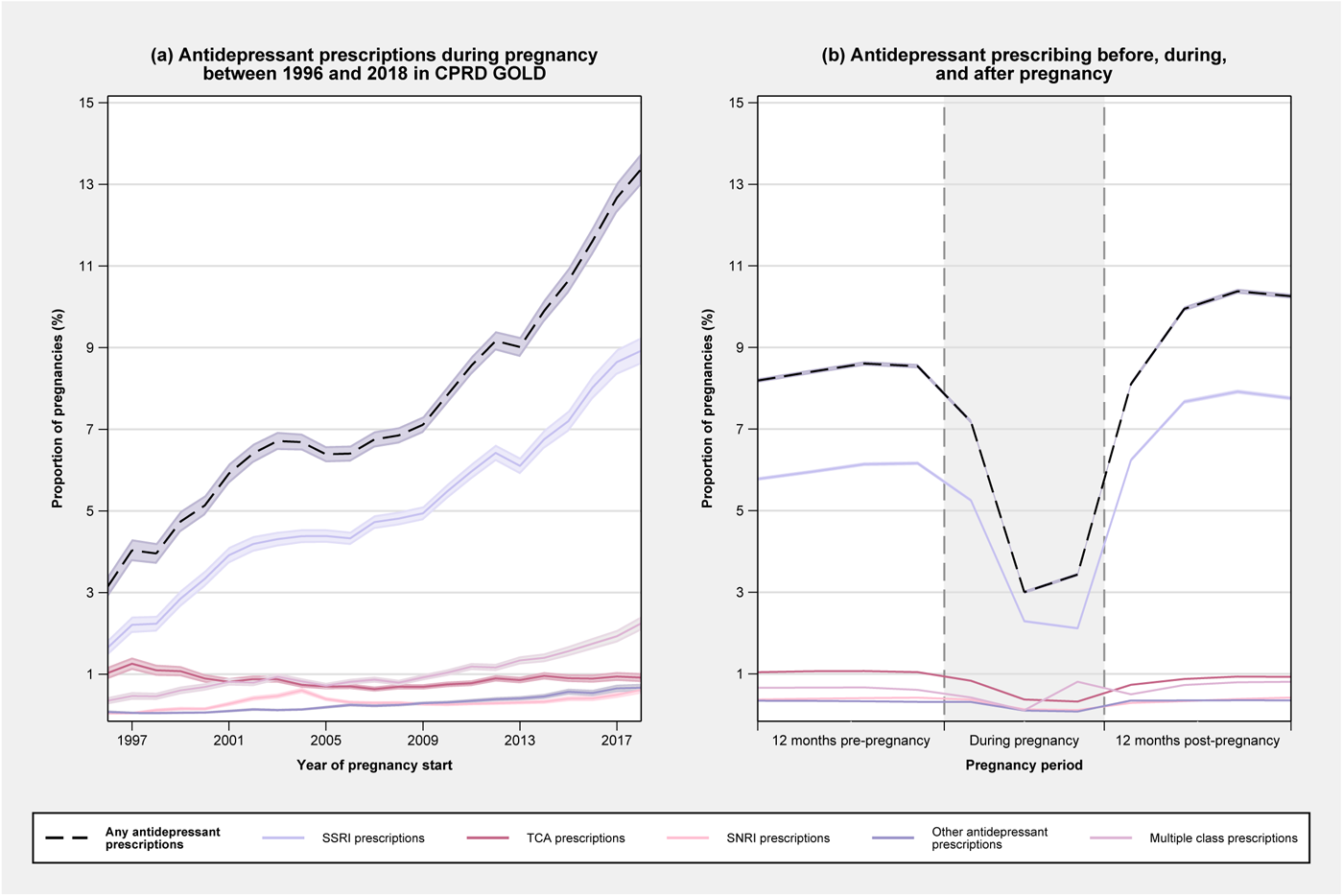
(a) Antidepressant prescribing during pregnancy over time in CPRD GOLD and (b) Proportion of pregnancies in the sample who were prescribed antidepressants before, during, and after pregnancy. The denominator for trimesters two and three consists of those whose pregnancies reached trimesters two and three, respectively, i.e., pregnancy losses in trimester one do not contribute to the denominator for trimesters two and three.

Wales had the highest overall rate of antidepressant prescribing during pregnancy (9.5% of all pregnancies during the study period, *n*=12,185) and London had the lowest rate (4.6% of all pregnancies during the study period, *n*=77,744) (Table S4).

### 3.3 Patterns of prescribing

#### 3.3.1 Pre-pregnancy

Of the 142,817 individuals prescribed antidepressants in the 12 months before pregnancy (13.8% of the study population), 92,670 discontinued prior to the start of pregnancy (64.9% of pre-pregnancy antidepressant users).

#### 3.3.2 During pregnancy

Of the 79,144 pregnancies among individuals who were prescribed antidepressants during pregnancy (7.7% of the study population) (Figure 1), 63,411 were ‘prevalent’ users (80.1% of antidepressant users during pregnancy). The remaining 15,733 (19.9%) were ‘incident’ users.

Most ‘prevalent’ users discontinued antidepressants during pregnancy (54.9%). Of the 42.8% who continued a single-drug regimen throughout pregnancy, the majority appeared to continue their regimen with no changes to dose or product (63.5%), 22.9% changed their dose, and 8.6% switched to a different product. The remaining 5.0% had evidence of multiple regimen changes during pregnancy (Table 2).

**Table 2.**
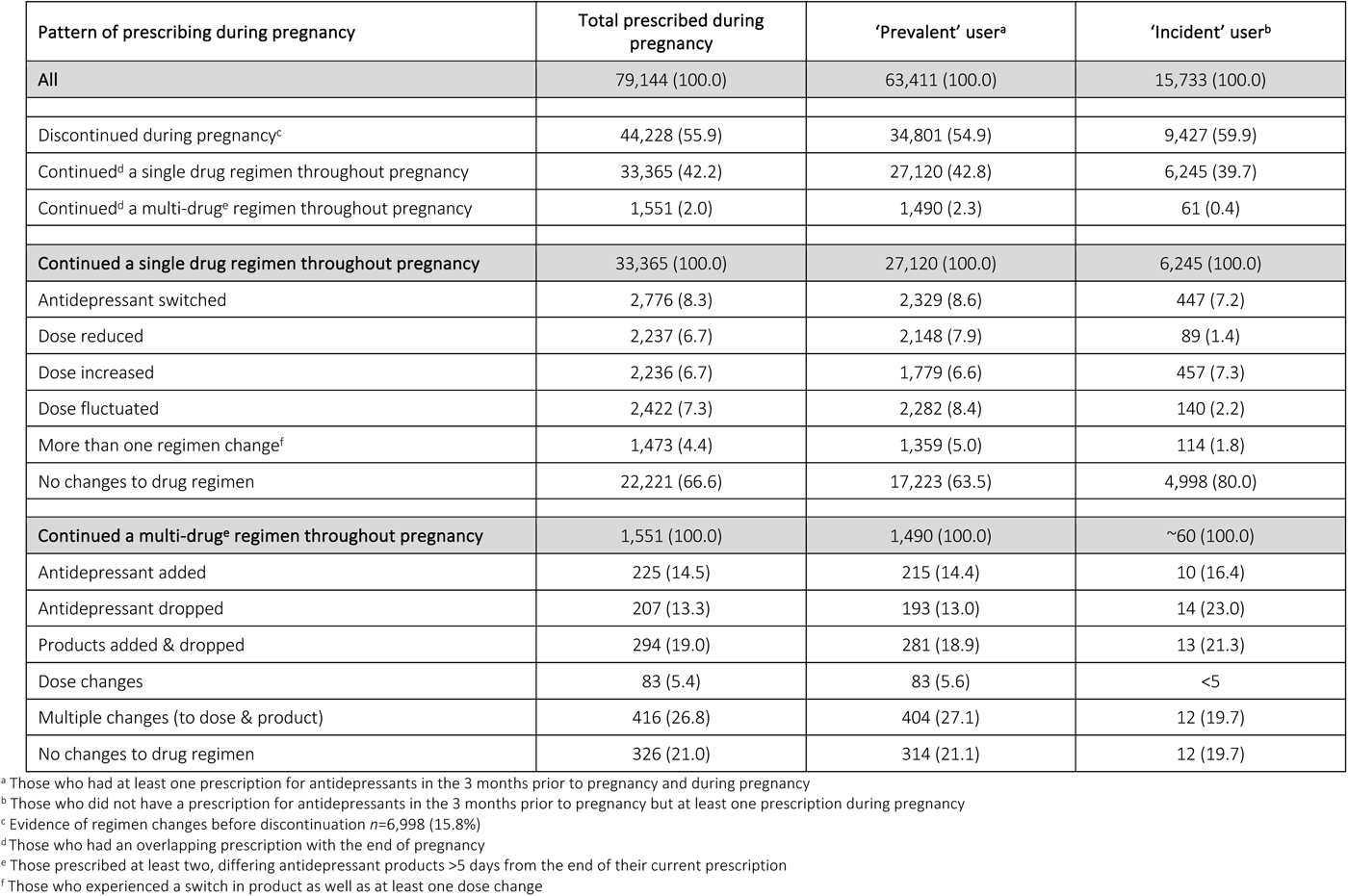
Patterns of prescribing during pregnancy.

| Pattern of prescribing during pregnancy | Total prescribed during pregnancy | 'Prevalent' user<sup>a</sup> | 'Incident' user<sup>b</sup> |
| --- | --- | --- | --- |
| **All** | 79,144 (100.0) | 63,411 (100.0) | 15,733 (100.0) |
| Discontinued during pregnancy<sup>c</sup> | 44,228 (55.9) | 34,801 (54.9) | 9,427 (59.9) |
| Continued<sup>d</sup> a single drug regimen throughout pregnancy | 33,365 (42.2) | 27,120 (42.8) | 6,245 (39.7) |
| Continued<sup>d</sup> a multi-drug<sup>e</sup> regimen throughout pregnancy | 1,551 (2.0) | 1,490 (2.3) | 61 (0.4) |
| **Continued a single drug regimen throughout pregnancy** | **33,365 (100.0)** | **27,120 (100.0)** | **6,245 (100.0)** |
| Antidepressant switched | 2,776 (8.3) | 2,329 (8.6) | 447 (7.2) |
| Dose reduced | 2,237 (6.7) | 2,148 (7.9) | 89 (1.4) |
| Dose increased | 2,236 (6.7) | 1,779 (6.6) | 457 (7.3) |
| Dose fluctuated | 2,422 (7.3) | 2,282 (8.4) | 140 (2.2) |
| More than one regimen change<sup>f</sup> | 1,473 (4.4) | 1,359 (5.0) | 114 (1.8) |
| No changes to drug regimen | 22,221 (66.6) | 17,223 (63.5) | 4,998 (80.0) |
| **Continued a multi-drug<sup>e</sup> regimen throughout pregnancy** | **1,551 (100.0)** | **1,490 (100.0)** | **~60 (100.0)** |
| Antidepressant added | 225 (14.5) | 215 (14.4) | 10 (16.4) |
| Antidepressant dropped | 207 (13.3) | 193 (13.0) | 14 (23.0) |
| Products added & dropped | 294 (19.0) | 281 (18.9) | 13 (21.3) |
| Dose changes | 83 (5.4) | 83 (5.6) | <5 |
| Multiple changes (to dose & product) | 416 (26.8) | 404 (27.1) | 12 (19.7) |
| No changes to drug regimen | 326 (21.0) | 314 (21.1) | 12 (19.7) |

<sup>a</sup> Those who had at least one prescription for antidepressants in the 3 months prior to pregnancy and during pregnancy
<sup>b</sup> Those who did not have a prescription for antidepressants in the 3 months prior to pregnancy but at least one prescription during pregnancy
<sup>c</sup> Evidence of regimen changes before discontinuation $n=6,998$ (15.8%)
<sup>d</sup> Those who had an overlapping prescription with the end of pregnancy
<sup>e</sup> Those prescribed at least two, differing antidepressant products >5 days from the end of their current prescription
<sup>f</sup> Those who experienced a switch in product as well as at least one dose change

Many ‘incident’ users also discontinued during pregnancy (59.9%). Of the ‘incident’ users who continued a single-drug regimen throughout pregnancy (39.7%), the majority did not make any changes to their regimen (80.0%). There was evidence of dose changes for a further 10.9%, drug switching in 7.2%, and multiple changes for 1.8% (Table 2).

When restricting to discontinuers with at least 29 completed weeks’ gestation, the majority discontinued in trimester one (77.7%). This was observed for both ‘prevalent’ and ‘incident’ users (81.8% and 63.4%, respectively) (Table S5).

We restricted the primary analysis to deliveries, then to losses (Table S3). The patterns of prescribing during pregnancy among deliveries was similar to the primary analysis, with 65.4% discontinuing during pregnancy and 54.5% of the single-drug continuers making no changes to their regimen (Table S6). Conversely, most women whose pregnancies ended in a loss continued antidepressants throughout pregnancy (62.8%), reflecting the shorter length of gestation (Table S7).

When restricting the sample to those with linked HES, the distribution of prescribing patterns was not altered in either ‘prevalent’ or ‘incident’ users (Table S8).

#### 3.3.3 Post-pregnancy

In the 12 months after pregnancy, 15.8% of the eligible sample received at least one prescription for antidepressants (*n*=162,947, Table S9), representing a slight increase from pre-pregnancy (**Error! Reference source not found**.).

Of the patients who discontinued within 12 months prior to pregnancy, 34.2% resumed antidepressant treatment in the 12 months after pregnancy (*n*=25,532). However, for those who discontinued during pregnancy, the majority resumed antidepressant treatment in the 12 months after pregnancy (53.0%, *n*=23,457) (Table S9).

When investigating post-pregnancy prescribing by delivery type, 58.2% of discontinuers during a pregnancy that ended in a loss resumed after pregnancy, compared to 51.5% of pregnancy discontinuers who had a delivery (Table S10). When restricting to those with at least 12 months of follow-up after the end of pregnancy, our conclusions did not change (Table S11).

### 3.4 Indications

Of those prescribed antidepressants during pregnancy, 80.6% had evidence of depression ever before the end of pregnancy (Table 1). Depression was first noted in 13.2% in the 12 months prior to pregnancy and 4.9% had incident antenatal depression of those prescribed antidepressants during pregnancy. Among the same group, 48.3% had evidence of anxiety before the end of pregnancy. Anxiety was first noted in the 12 months before pregnancy in 7.5% of women prescribed antidepressants during pregnancy, and first noted during pregnancy in 3.0% among those prescribed antidepressants during pregnancy (Table S12).

Among those prescribed antidepressants during pregnancy, 9.6% had no evidence of an antidepressant indication (Table S13).

### 3.5 Predictors of discontinuation

Younger (<18 years OR 1.41 95%CI 1.24–1.60) and 18-24 years OR 1.37 95%CI 1.32–1.43), underweight (OR 1.25 95%CI 1.15–1.36), and more deprived (most deprived OR 1.31 95%CI 1.24–1.37) women were more likely to discontinue antidepressants than comparators. Current and ex-smokers (OR 0.93 95%CI 0.90–0.97 and OR 0.91 95%CI 0.88–0.94, respectively) were less likely to discontinue than non-smokers and those who had experienced a stillbirth (OR 1.19 95%CI 1.01–1.40) were more likely to discontinue than those who hadn’t. Those with higher parity (≥3 OR 0.71 95%CI 0.67–0.74) and previous miscarriage (OR 0.90 95%CI 0.87–0.93) were less likely to discontinue during pregnancy (Figure 2).

**Figure 2.**
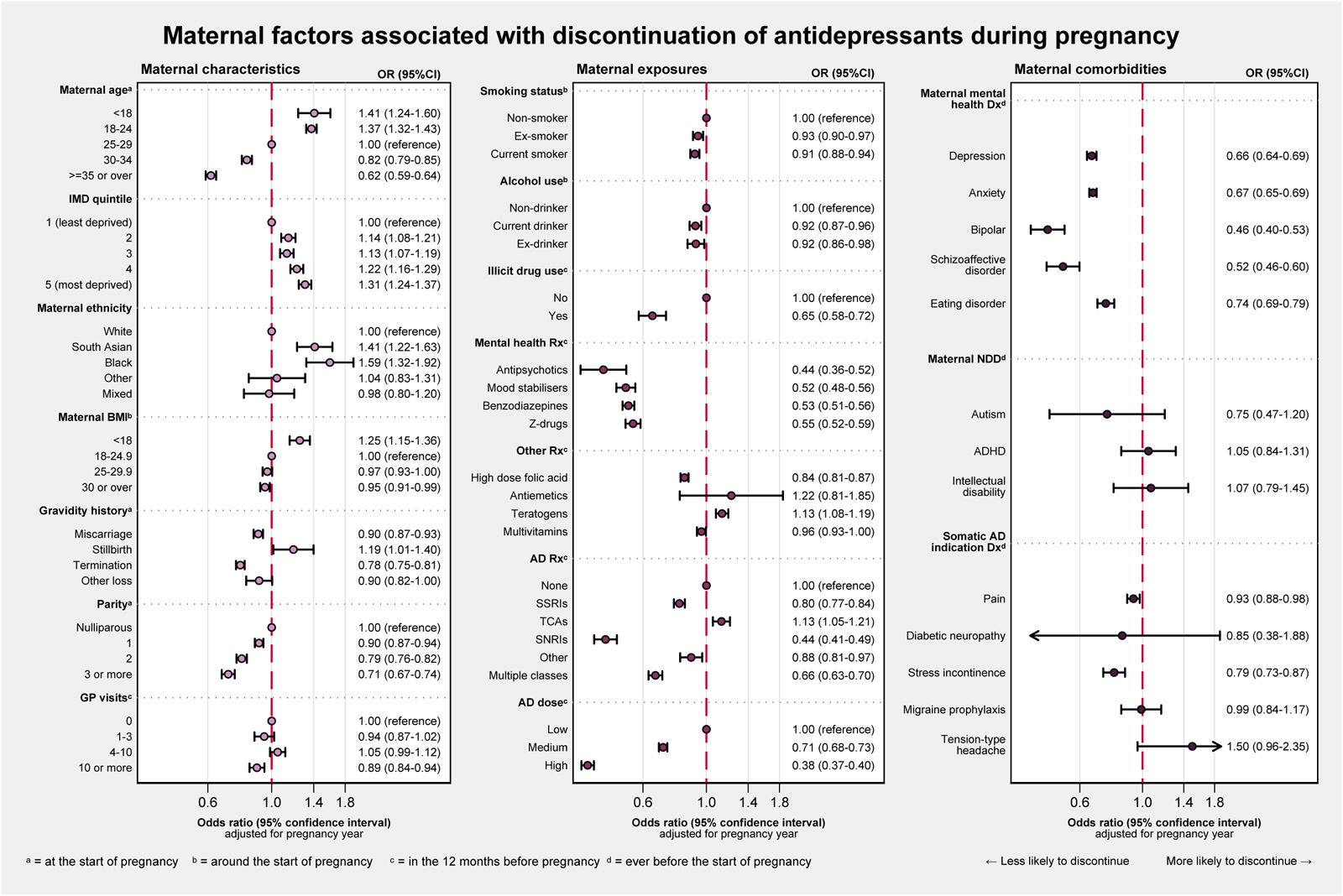
Maternal factors associated with discontinuation of antidepressants during pregnancy. Rx = prescription Dx = evidence of a diagnosis

Ethnicity, BMI, smoking, and alcohol use around the start of pregnancy had varying degrees of missing data. Discontinuing during pregnancy increased the likelihood of having missing data in BMI, smoking, and alcohol use as compared to continuing throughout pregnancy (Table S14).

## Discussion

### Main findings

The present study gives a detailed overview of antidepressant prescribing in and around pregnancy in the UK between 1996 and 2018, highlighting an increase from 3.2% in 1996 to 13.4% in 2018. Utilising all prescriptions made during pregnancy, we describe patterns of antidepressant prescribing during pregnancy and show the high resumption rate soon after the end of pregnancy among those who discontinued during pregnancy (53.0%).

Of both ‘prevalent’ and ‘incident’ users of antidepressants during pregnancy, the majority discontinued their regimen at some point during pregnancy (54.9% and 59.9%, respectively). Most measured demographics were associated with discontinuation: younger age, underweight BMI, lower SEP, and history of stillbirth were associated with an increased likelihood of discontinuation. Whereas, older maternal age, pre-pregnancy polypharmacy, and being a smoker was associated with a decreased likelihood of discontinuing. Of the ‘prevalent’ users who continued their regimen throughout pregnancy, 51% continued their regimen with no regimen changes, as opposed to over 70% of ‘incident’ users.

Those prescribed antidepressants during pregnancy have been shown to have greater needs and require more support during pregnancy.^28^ It is important to understand this group of people to provide the best care in and around pregnancy.

### Strengths and limitations

The present study has several strengths. It is a large, population-based study that includes pregnancies of all known outcomes, not restricted to live birth, using a validated pregnancy register linked to primary and secondary care.^23^ It is the first study to discuss antidepressant prescribing alongside indication prevalence in and around the pregnancy period. Given that antidepressants are not sold over the counter in the UK and mostly prescribed in primary rather than secondary care, we are confident that we captured the majority of antidepressant prescribing in the eligible population.

The study does however have several limitations. The number of pregnancies has been dropping in CPRD GOLD in the last decade, likely due to more women self-referring to a midwife and circumventing her GP.^29^ It is possible that women on antidepressants are more likely to report their pregnancy to the GP than women who are not, which would artificially inflate the proportion of women on antidepressants during pregnancy after 2010 in the eligible sample and make the increase in antidepressant prescribing during pregnancy look greater than is true in the general population. However, antidepressant prescribing during pregnancy increased from 3.2% in 1996 to 7.1% in 2009 before these changes were enacted, reflecting trends outside of pregnancy among women of a child-bearing age.

Despite updating unknown outcome pregnancies and conflicts where possible,^24^ 9% of pregnancies were unresolved and thus dropped. This may have introduced selection bias and we may have incorrectly estimated the prevalence of certain prescribing practices or the associations between different demographics and discontinuation. Although gradual dose reduction is recommended when discontinuing antidepressants,^30^ there was limited evidence of this in the prescription data. However, it is plausible that dose reductions may have been described by the prescriber in the free text, that were then missed in the available structured data fields.

Pregnancy length was imputed for some pregnancies in the Pregnancy Register; this is more common for losses where less information on the pregnancy is available, potentially putting the study at risk of differential antidepressant exposure misclassification. We may have been more likely to misclassify losses as antidepressant prescribed when they weren’t, thus have overestimated antidepressant exposure and certain patterns among this group. We used prescriptions of antidepressants to proxy exposure, but we had no information on dispensation or adherence, so some individuals may have been misclassified if they never filled or took their prescription. Identifying those on a multi-drug regimen was challenging; it was difficult to differentiate an antidepressant switch from a multi-drug regimen in many cases and some multi-drug regimens may have been misclassified as product switching.

Missing data was a problem in some of the covariates, such as smoking and BMI. In sensitivity analysis, we observed an association between missingness in ethnicity, BMI, smoking, and alcohol use and an increased likelihood to discontinue antidepressants, suggesting there may be a risk of bias in the CRA for these characteristics,^27^ so should be interpreted with caution.

### Interpretation

Our estimate of antidepressant prescribing during pregnancy is in line with the trajectory identified by Petersen *et al.* in 2011, who reported a 4-fold increase in antidepressant prescribing during pregnancies that ended in live birth between 1992 and 2006 in the UK.^31^ The upwards trend over time reflects the increased antidepressant prescribing in the general UK population over recent decades^17, 32^ and shows the growing need for evidence-based advice on antidepressant use during pregnancy. Most individuals who were using antidepressants during pregnancy discontinued, predominantly in trimester one. NICE guidance notes that antidepressants can be used at any stage of pregnancy if clinically indicated, but that their risks and benefits should be specific to each woman.^5–7^ However, the evidence regarding risks and the efficacy of these guidelines in reducing them, is mixed or unknown.

In relation to patterns of prescribing, the findings were in line with previous literature.^19, 21^ We found that continuation without dose changes was more common among ‘incident’ than ‘prevalent’ users, due to the likelihood that clinically, ‘incident’ users would be initiated on and maintain a low dose if symptoms were managed.

It is important to note the high post-pregnancy antidepressant resumption rate among those who discontinued antidepressants during pregnancy, which remained high when stratified by delivery and pregnancy loss (51.5% and 58.2%, respectively). A small study from France identified both benzodiazepine and anxiolytic use after pregnancy was higher than pre-pregnancy among those who discontinued antidepressants during pregnancy suggesting that symptoms may worsen when interrupting treatment.^33^ High antidepressant resumption rate may potentially reflect an exacerbation of illness during or after pregnancy.

The importance of descriptive epidemiology, here in the context of drug utilisation, cannot be underestimated.^34^ It underpins subsequent studies aimed to assess causality in an observational setting by highlighting important demographics among exposure groups of interest, key differences between them and potential comparator groups, and data pitfalls that might hinder causal inference. The present study provides a useful resource for both researchers hoping to contribute high-quality evidence regarding the safety of antidepressant use during pregnancy and clinicians who are interested in the trends of different prescribing patterns in and around pregnancy.

## Conclusion

Antidepressant use during pregnancy increased between 1996 and 2018 in the UK, from 3.2% to 13.4%. The majority (55.9%) of individuals prescribed antidepressants during pregnancy discontinued at some point before the end of pregnancy; resumption rate in the 12 months after pregnancy was high (53.0%) among these individuals. Future studies would be useful to ascertain the impact of different antidepressant prescribing patterns on maternal and fetal health to better inform clinical guidance and practice.

## Data Availability

All data produced in the present work are contained in the manuscript.

## Author contributions

FZM, DR, GCS, HF, and KEE proposed the original study, and FM provided the initial draft of the study. DR, VNS, and AS assisted with the clinical sign-off for the codelists; JLR provided expertise in codelist creation also. DR, LB, VNS, and AS provided clinical and topical expertise and interpretation of the findings. GCS, KEE, LB, PMD, JLR, DR, and HR provided epidemiological expertise. PMD, HF and DR contributed methodological and data expertise to the design and write-up of the study. FZM performed the data analysis. All authors contributed to the preparation and editing of the manuscript and approved the final paper for submission.

## Acknowledgements

Thank you to Neil M Davies and his team for their support with data acquisition. Thanks to Abigail Merriel for her help with assessing clinical codelists.

