## Supplementary material for "Patterns of antidepressant prescribing in and around pregnancy: a descriptive analysis in the UK Clinical Practice Research Datalink": Methods S, Table S, Figure S

### 1 METHODS

#### 1.1 DATA SOURCES

CPRD GOLD is an ongoing resource that collects data from consenting primary care sites in the UK that use a specific computer software system called Vision.[1] It is one of the largest primary care datasets in the world, containing over 4 million active patients (around 7% of the whole UK population) that have been shown to be representative of UK residents in terms of age, sex, and ethnicity.[1] Over 670 practices in England, Scotland, Wales, and Northern Ireland have consented to practice- and patient-level data abstraction by CPRD, where each patient has data available on prescriptions (using British National Formulary (BNF) codes), diagnoses (using Read codes), and characteristics collected since their registration date at the practice.[1]

We used linked data, where available. Linkage with the Office for National Statistics (ONS, capturing death registrations) and practice-level Index for Multiple Deprivation scores (IMD, capturing patient and practice-level deprivation using scores predefined by geographical area)[2] are available for all patients in CPRD GOLD. Other sources of linked data, including patient-level IMD scores, those from Hospital Episode Statistics (HES, capturing inpatient hospitalisations) and the Mental Health Services Data Set (MHSDS, capturing inpatient and outpatient contacts with mental health services),[1, 3] are only available for 75% of English practices.[1]

The Pregnancy Register contains all pregnancies for women of child-bearing age in CPRD GOLD, including estimated timings of pregnancy (from multiple dates recorded throughout pregnancy), pregnancy outcome (such as live birth and miscarriage), and maternal age.[4] Close agreement with hospitalization data show that the data included within the Pregnancy Register are well recorded.[4]

A GP is often the first medical professional involved in providing antenatal care; the NHS website recommends that individuals make an appointment with their GP as soon as they know they are pregnant; the GP can then coordinate further appointments with midwifery and obstetric services.[5] However since 2007, women have had the choice between registering their pregnancy with their GP or going directly to register with a midwife.[6] Although notes should be reported back to the GP if the latter occurs, it may not be consistent and may explain the decline in number of pregnancies in CPRD GOLD from around 2010.[6]

#### 1.2 Identifying pregnancies

Individuals were dropped from the analysis if they did not have at least 12 months of follow-up with an 'up to standard' (UTS) practice. Practices are assigned an UTS date which refers to the date on which the practice started contributing 'high-quality' data and is used in CPRD studies as an internal quality control.[1]

The Pregnancy Register as received from CPRD contains the outcome of the pregnancy (livebirth, stillbirth, miscarriage, etc.). A substantial proportion of the outcome variable consists of "Unspecified loss" and "Outcome unknown" pregnancies, detailed elsewhere.[4] Given that the Pregnancy Register algorithm only used primary care data to identify pregnancy episodes and their outcome, we employed an additional approach to try and convert unspecified outcomes to know outcomes using secondary care data. If a patient received care in hospital for the resolution of their pregnancy (either miscarriage care or a delivery), we could identify records in HES that are likely to pertain to pregnancy episodes in the Pregnancy Register to recode unknown outcomes to known.

Using the algorithmic approach described by Campbell *et al.*[7], we pulled together episodes from secondary care pertaining to pregnancy where the outcome was known and then aligned the dates with the estimated dates of the pregnancy episode in the Pregnancy Register. In doing so, we could use a combination of the recorded outcome and episode date in HES in place of the unknown outcome and estimated pregnancy end date in the Pregnancy Register, with the imputed gestational length employed by the Pregnancy Register algorithm to estimate an updated pregnancy start (subtracting the imputed length from the episode date in HES).

#### 1.3 ANTIDEPRESSANT PRESCRIBING

Given that prescriptions for antidepressants are made in primary care, prescription timing and estimated length of prescription were used in this study to ascertain whether pregnant patients may have been exposed to antidepressants during pregnancy and how their regimens may have changed over the course of pregnancy.

Within CPRD, multiple variables are derived from dosage text, inputted by the GP into the Vision software, namely dose number, dose frequency, and daily dose. Dose number corresponds to the number of doses taken each time the medicine is advised, and dose frequency refers to the number of times a dose should be administered daily; thus, for "2 tablets to be taken twice daily" the dose number would be two and the dose frequency would be two. Daily dose is the dose number multiplied by the dose frequency, in other words the total number of doses taken each day, so for the above example the daily dose would be four.

Dose number and dose frequency were cleaned to obtain any additional information from dosage text and used to derive a clean daily dose for each prescription. Abnormal values for dose number (0 and >15) were changed to missing so that daily dose could be imputed in a later step. To ascertain each prescriptions length, quantity was also cleaned (the total number of dose unit given for each prescription) to combine with daily dose. Similarly to dose number, abnormal values (< 5 or > 300) were changed to missing and imputed later.

For missing values in daily dose and quantity, a hot-decking imputation approach was used, which employed a stepwise process to impute the modal daily dose and quantity at decreasing levels of specificity. Firstly, modal values were taken from identical prescriptions made in the same person if not missing. If still missing, modal values were taken from identical prescriptions in different people. If still missing, modal values were taken from the same medication prescribed at different quantities/daily doses. This decreasing specificity fills in the last few missing values with modal values of medications in that wider class. Following the hot-decking imputation step, these complete data that contained prescription start and end dates was merged with the Pregnancy Register to align with complementary pregnancy start and end dates. We were then able to identify prescriptions that fell within each period of interest, meaning individuals could be categorised as exposed (having received a prescription) or unexposed (having not received a prescription) during each period (before, during, and after pregnancy).

To identify dose changes, we standardised doses across each medication so they could be compared. We employed an approach that leveraged the distribution of daily doses (in milligrams) within each drug in the data, using the quartile values to assign low, medium, and high. For example, fluvoxamine had a range of 25–600mg per day in our data. We then assigned the lowest 25% of the daily dose "low" (≤25$^{th}$ percentile, 25–50mg), the middle 50% "medium" (>25$^{th}$ and ≤75$^{th}$ percentile, 75–100mg), and the upper quartile "high" (>75$^{th}$ percentile, 112.5–600mg). Some rarely prescribed medications had small distributions due to lack of diversity in the doses prescribed, meaning that the same prescribed dose was accounted for in the quartiles and more stringent thresholds were required. For example, the isocarboxazid distribution contained 20mg in the 25$^{th}$, 50$^{th}$, and 75$^{th}$ quartile so the same thresholds were applied as above but with the 25$^{th}$, 75$^{th}$, and 90$^{th}$ percentile. For isocarboxazid, “low” pertained to 10–20mg, “medium” to 22.5mg, and “high” to 30–40mg. These were checked against the EMC website to confirm that they aligned with clinical practice. The summary of each dose assignment for each drug is provided as an Excel spreadsheet with the supplementary material.

Upon inspecting the data, some prescriptions retained needed some additional cleaning. There were prescriptions for the same product that had been made on the same day for the same duration but had different affiliated daily doses. Following clinical input from medical co-authors, these were deemed to be the same prescription where the prescribing clinician was creating a dose amount not available as a singular tablet (for example, to achieve a 30mg dose of citalopram, the prescription would appear across three records: a 20mg and a 10mg). For these prescriptions where the product, prescription length, and prescriptions dates were the same, but the dose was different for each, the daily doses in milligrams were added together and “duplicates” were removed. The above process of assigning standardised doses was then reapplied to assure that the new combined doses had been given the right category: low, medium, or high.

**Table S1** Drug substances that fall into each class category of antidepressants: SSRIs, SNRIs, TCAs, and 'other'.

| Selective serotonin reuptake inhibitors (SSRIs) | (Serotonin-) noradrenaline reuptake inhibitors (SNRI/NRIs) | Tricyclic antidepressants (TCAs) | 'Other' antidepressants |
|---|---|---|---|
| Citalopram | Duloxetine | Amitriptyline | Agomelatine |
| Escitalopram | Reboxetine | Amoxapine | Isocarboxazid |
| Fluoxetine | Venlafaxine | Clomipramine | Mianserin |
| Fluvoxamine | | Desipramine | Mirtazapine |
| Paroxetine | | Dosulepin | Nefazodone |
| Sertraline | | Doxepin | Phenelzine |
| | | Imipramine | Tranylcypromine |
| | | Lofepramine | Trazodone |
| | | Maprotiline | Tryptophan |
| | | Nortriptyline | Vortioxetine |
| | | Protriptyline | |
| | | Trimipramine | |

#### 1.4 Patterns of prescribing

During pregnancy, discontinuation was reported as any during pregnancy and was defined as the final antidepressant prescription ending more than 2 weeks from the end of pregnancy. In sensitivity analysis, trimester of discontinuation was explored among those with at least 27 completed weeks' gestation: trimester one (final prescription during pregnancy ending in between week 14), trimester two (final prescription during pregnancy ending in between week 14 and week 27), and trimester three. Trimester three discontinuation was defined by those with a prescription for antidepressants during trimester three but no prescriptions for antidepressants in the 3 months after the end of pregnancy (Figure S1).

We differentiated between single- and multi-drug regimens (mono- and polytherapy) for those who did not discontinue during pregnancy. Those with two prescriptions for different antidepressant products overlapping by more than four weeks were considered to be using a multi-drug regimen (to prevent cross-tapering[8] being wrongly considered as a multi-drug regimen, as opposed to switching within a single-drug regimen), whereas those that did not were classified as individuals prescribed a single-drug regimen.

Among those on a single-drug regimen, we showed product switching (subsequent use of a different antidepressant) and dose changes (increase or decrease with the same product). For patients on a multi-drug regimen, we described product adding to and dropping from the regimen, as well as dose changes within products in the regimen. Example prescription patterns that fulfil the criteria for each pattern are shown in Figure S2.

**Figure S1** Type of antidepressant use before and during pregnancy.

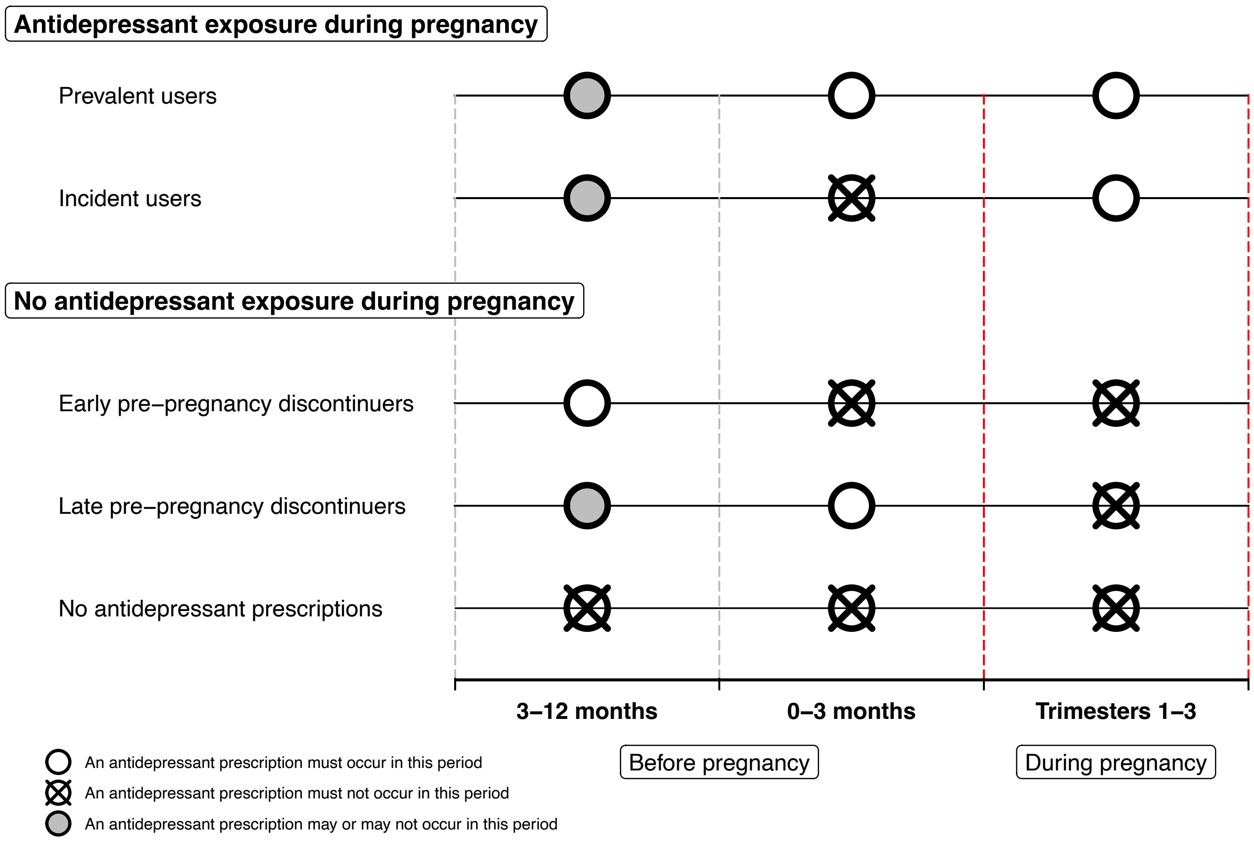

**Figure S2** Examples of pattern definition for those prescribed to antidepressants during pregnancy.

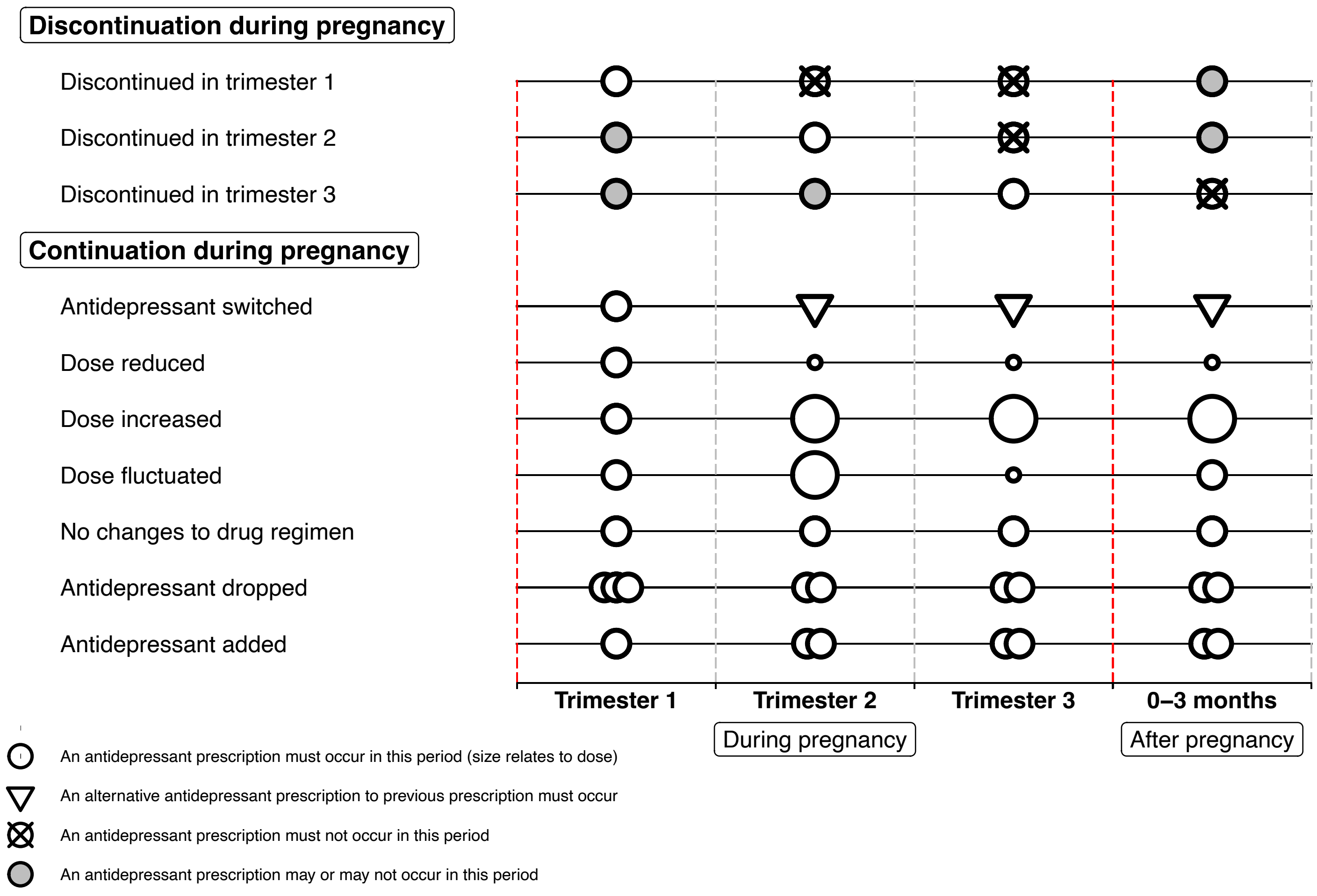

#### 1.5 CHARACTERISTIC DERIVATION

Indication-based lists of Read code were created using the medical dictionaries provided by CPRD. ICD-10 codelists were then created using keyword searches. Both were checked by clinical co-authors and are available in github/patterns. The Read code lists were then faced to the Clinical and Referral data in CPRD GOLD, and the ICD-10 code lists were faced towards the linked HES data for whom it was available.

Other characteristics were obtained from CPRD and linked data and summarised in Table S2.

**Table S2** Details of covariate definitions.

| Covariate | Definition |
|---|---|
| Maternal age at start of pregnancy | Given that only year of birth available in CPRD for anonymisation, a pseudo-date of birth of 1st July in each patient's year of birth is generated and then subtracted from the pregnancy start date |
| Practice IMD (in quintiles) | The Index of Multiple Deprivation (IMD) is a composite measure of several domains that capture 'deprivation': a score is attributed to each practice ID and then all scores are divided into quintiles, with quintile 1 being least deprived and quintile 5 being most deprived |
| Maternal ethnicity | Maternal ethnicity is obtained from both CPRD Clinical data and Hospital Episode Statistics (HES) Patient data |
| Maternal smoking | Maternal smoking during pregnancy, recorded in Clinical and Additional data, if available; if not the algorithm takes the next nearest record of smoking within 10 years of pregnancy start |
| Maternal alcohol intake | Maternal alcohol intake during pregnancy, recorded in Clinical and Additional data, as well as pharmacological treatments for alcohol dependence in Therapy data, if available; if not the algorithm takes the next nearest record of alcohol intake within 10 years of pregnancy start |
| Maternal illicit drug use | Evidence of illicit drug use any time during pregnancy in the Clinical or Referral data or prescriptions for illicit drug use e.g., methadone any time during pregnancy in the Therapy data |
| Maternal gravidity history at start of pregnancy | History of miscarriage, stillbirth, termination of pregnancy, or other losses (ectopic pregnancy, molar pregnancy, blighted ovum etc.) at the start of pregnancy |

| | |
|---|---|
| Maternal parity at start of pregnancy | Number of pregnancies carried to more than 20 weeks' gestation (live births, stillbirths, deliveries based on third trimester or late pregnancy records) at the start of pregnancy |
| Number of consultations in CPRD in the 12 months before pregnancy | The number of consultations each patient had in the year prior to pregnancy derived from the CPRD Consultation data |
| Number of consultations in CPRD during pregnancy | The number of consultations each patient during pregnancy derived from the CPRD Consultation data |
| Maternal mental health problems ever before or during pregnancy | Evidence of the first diagnosis of mental health disorders e.g., depression, anxiety, bipolar etc. ever before or during pregnancy (i.e., diagnosis made prior to the pregnancy end date) in the Clinical or Referral data, as well as HES for whom it was available |
| Maternal neurodevelopmental disorders ever before or during pregnancy | Evidence of the first diagnosis of neurodevelopmental disorders e.g., autism spectrum disorder, attention deficit hyperactivity disorder etc. ever before or during pregnancy (i.e., diagnosis made prior to the pregnancy end date) in the Clinical or Referral data |
| Possible somatic indications for antidepressants ever or during pregnancy | Evidence of the first diagnosis of other possible somatic indications for antidepressants e.g., pain, tension-type headache, migraine prophylaxis etc. ever before or during pregnancy (i.e., diagnosis made prior to the pregnancy end date) in the Clinical or Referral data |
| Other mental health-related prescriptions during pregnancy | Mental health-related prescriptions including mood stabilisers, benzodiazepines, antipsychotics etc. made during pregnancy identified from the Therapy data |
| Other prescriptions during pregnancy | Other prescriptions including anti-emetics, folic acid, multivitamins etc. made during pregnancy identified from the Therapy data |

### 2 RESULTS

#### 2.1 STUDY POPULATION

**Figure S3** Detailed flow of pregnancies through the study.

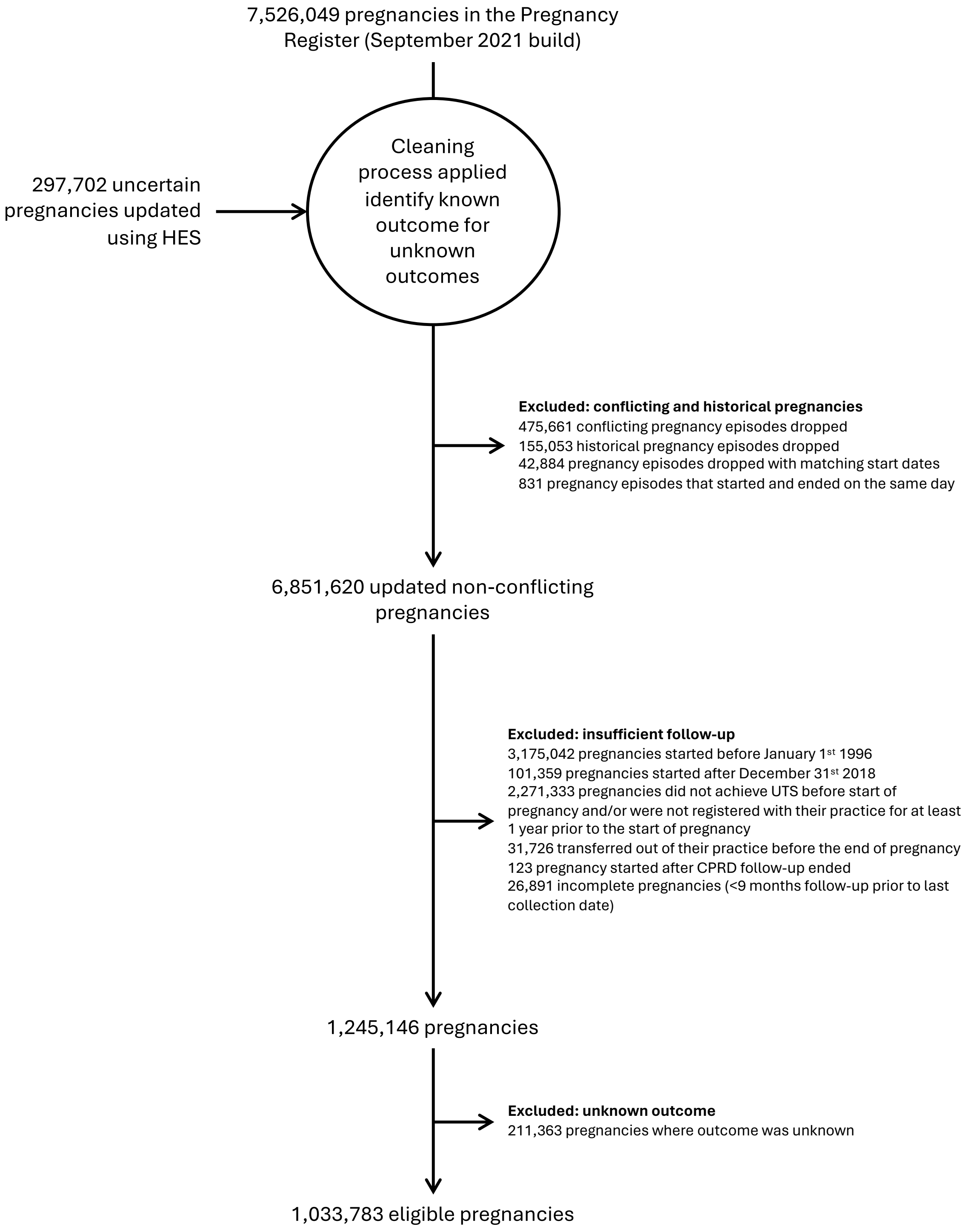

**Table S3** Proportion of pregnancy outcomes in eligible sample

| Outcome | Proportion | Delivery or loss |
|---|---|---|
| Live birth | 734,938 (71.09) | Delivery |
| Stillbirth | 3,211 (0.31) | Delivery |
| Live birth or stillbirth | 549 (0.05) | Delivery |
| Miscarriage | 127,521 (12.34) | Loss |
| Termination of pregnancy (TOP) | 39,744 (3.84) | Loss |
| Probable TOP | 99,841 (9.66) | Loss |
| Ectopic | 11,158 (1.08) | Loss |
| Molar | 975 (0.09) | Loss |
| Blighted ovum | 738 (0.07) | Loss |
| Unspecified loss | 6,495 (0.63) | Loss |
| Delivery based on a third trimester record | 6,640 (0.64) | Delivery |
| Delivery based on a late pregnancy record | 1,973 (0.19) | Delivery |

#### 2.2 Prescribing over time among live births

**Figure S4** Antidepressant prescribing during pregnancy restricted to live births.

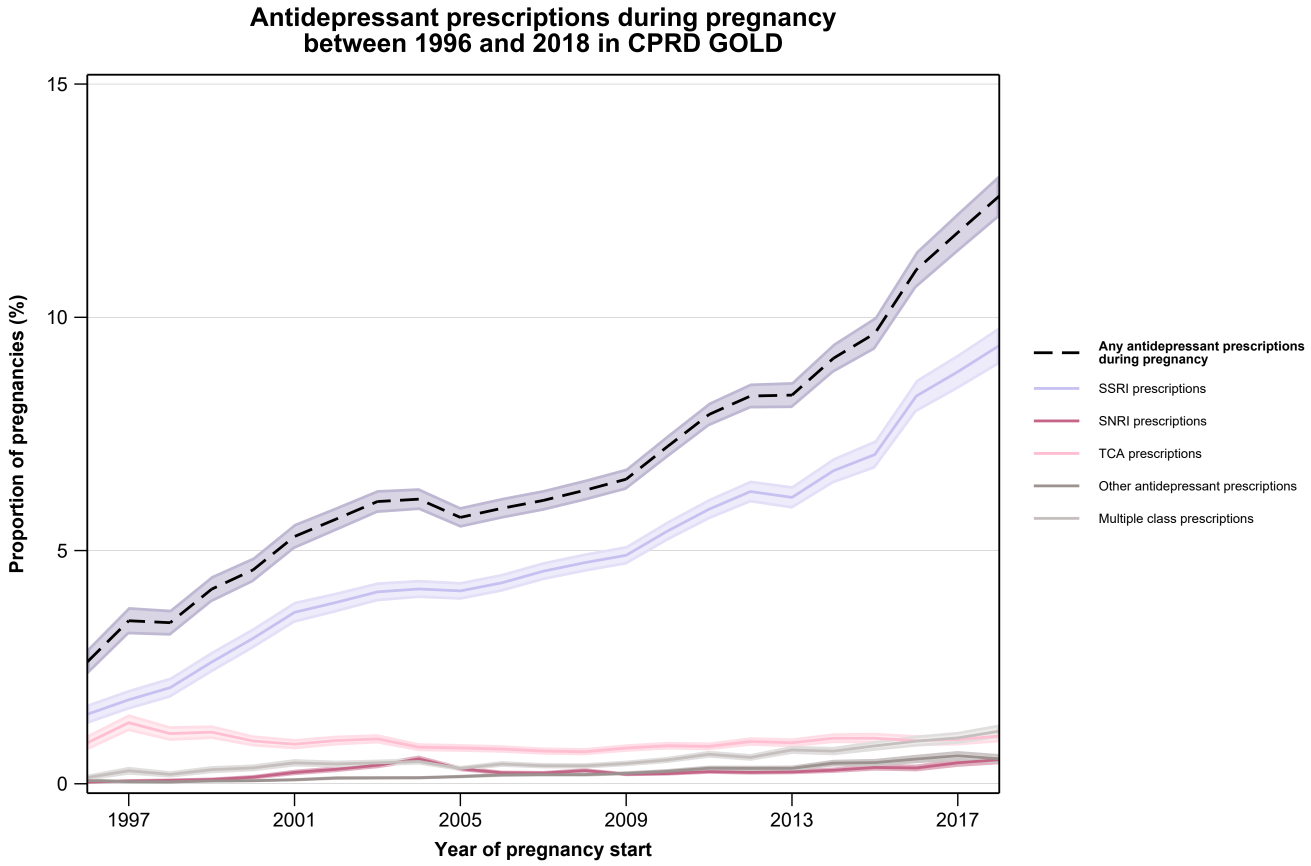

#### 2.3 PRESCRIBING BY REGION

**Table S4** Proportion of pregnancies from each CPRD region prescribed antidepressants during pregnancy.

| Practice region | Prescribed/Total | % |
|---|---|---|
| East Midlands | 1,832/27,468 | 6.67 |
| East of England | 4,228/66,939 | 6.32 |
| London | 3,606/77,744 | 4.64 |
| North East | 1,040/15,932 | 6.53 |
| North West | 8,216/110,062 | 7.46 |
| Northern Ireland | 5,010/53,654 | 9.34 |
| Scotland | 19,056/204,963 | 9.30 |
| South Central | 6,030/86,163 | 7.00 |
| South East Coast | 6,062/86,510 | 7.01 |
| South West | 5,040/67,612 | 7.45 |
| Wales | 12,185/128,681 | 9.47 |
| East Midlands | 5,348/81,697 | 6.55 |
| Yorkshire & The Humber | 1,491/26,358 | 5.66 |

#### 2.4 DISCONTINUATION BY TRIMESTER

**Table S5** Patterns of discontinuation among those with at least 27 completed weeks' gestation.

| Pattern of discontinuation during pregnancy | Total prescribed during pregnancy[a] | 'Prevalent' user[b] | 'Incident' user[c] |
|---|---|---|---|
| All | 33,988 (100.0) | 26,441 (100.0) | 7,547 (100.0) |
| Discontinuation in trimester 1[d] | 26,422 (77.7) | 21,637 (81.8) | 4,785 (63.4) |
| Discontinuation in trimester 2[e] | 5,465 (16.1) | 3,713 (14.0) | 1,752 (23.2) |
| Discontinuation in trimester 3[f] | 2,101 (6.2) | 1,091 (4.1) | 1,010 (13.4) |

[a] Restricted to those with at least 27 completed weeks' gestation
[b] Those who had at least one prescription for antidepressants in the 3 months prior to pregnancy and during pregnancy
[c] Those who did not have a prescription for antidepressants in the 3 months prior to pregnancy but at least one prescription during pregnancy
[d] Evidence of regimen changes before discontinuation *n*=3,465 (13.1%)
[e] Evidence of regimen changes before discontinuation *n*=1,854 (33.9%)
[f] Evidence of regimen changes before discontinuation *n*=680 (32.4%)

#### 2.5 Patterns among deliveries

**Table S6** Patterns analysis restricted to delivery outcomes (live birth, stillbirth, live birth or stillbirth, delivery based on a third trimester record, delivery based on a late record).

| Pattern of prescribing during pregnancy | Total deliveries prescribed during pregnancy | 'Prevalent' user[b] | 'Incident' user[c] |
|---|---|---|---|
| **All** | 52,306 (100.0) | 41,635 (100.0) | 10,671 (100.0) |
| Discontinued during pregnancy[c] | 34,234 (65.4) | 26,638 (64.0) | 7,596 (71.2) |
| Continued[d] a single drug regimen throughout pregnancy | 17,072 (32.6) | 14,043 (33.7) | 3,029 (28.4) |
| Continued[d] a multi-drug[e] regimen throughout pregnancy | 1,000 (1.9) | 954 (2.3) | 46 (0.4) |
| **Continued a single drug regimen throughout pregnancy** | 17,072 (100.0) | 14,043 (100.0) | 3,029 (100.0) |
| Antidepressant switched | 1,858 (10.9) | 1,571 (11.2) | 287 (9.5) |
| Dose reduced | 1,603 (9.4) | 1,540 (11.0) | 63 (2.1) |
| Dose increased | 1,104 (6.5) | 824 (5.9) | 280 (9.2) |
| Dose fluctuated | 2,009 (11.8) | 1,896 (13.5) | 113 (3.7) |
| More than one regimen change[f] | 1,193 (7.0) | 1,101 (7.8) | 92 (3.0) |
| No changes to drug regimen | 9,305 (54.5) | 7,111 (50.6) | 2,194 (72.4) |
| **Continued a multi-drug[e] regimen throughout pregnancy** | 1,000 (100.0) | 954 (100.0) | 46 (100.0) |
| Antidepressant added | 128 (12.8) | 122 (12.8) | 6 (13.0) |
| Antidepressant dropped | 125 (12.5) | 116 (12.2) | 9 (19.6) |
| Products added & dropped | 210 (21.0) | 199 (20.9) | 11 (23.9) |
| Dose changes | 57 (5.7) | 57 (6.0) | <5 |
| Multiple changes (to dose & product) | 329 (32.9) | 317 (33.2) | 12 (26.1) |
| No changes to drug regimen | 326 (21.0) | 314 (21.1) | 12 (19.7) |

[a] Those who had at least one prescription for antidepressants in the 3 months prior to pregnancy and during pregnancy
[b] Those who did not have a prescription for antidepressants in the 3 months prior to pregnancy but at least one prescription during pregnancy
[c] Evidence of regimen changes before discontinuation *n*=6,041 (17.6%)
[d] Those who had an overlapping prescription with the end of pregnancy

[e] Those prescribed at least two, differing antidepressant products >5 days from the end of their current prescription
[f] Those who experienced a switch in product as well as at least one dose change

#### 2.6 Patterns among losses

**Table S7** Patterns analysis restricted to loss outcomes (miscarriage, TOP, probable TOP, ectopic, molar, blighted ovum, unspecified loss).

| Pattern of prescribing during pregnancy | Total losses prescribed during pregnancy | 'Prevalent' user[b] | 'Incident' user[c] |
|---|---|---|---|
| **All** | 26,838 (100.0) | 21,776 (100.0) | 5,062 (100.0) |
| Discontinued during pregnancy[c] | 9,994 (37.2) | 8,163 (37.5) | 1,831 (36.2) |
| Continued[d] a single drug regimen throughout pregnancy | 16,293 (60.7) | 13,077 (60.1) | 3,216 (63.5) |
| Continued[d] a multi-drug[e] regimen throughout pregnancy | 551 (2.1) | 536 (2.5) | 15 (0.3) |
| **Continued a single drug regimen throughout pregnancy** | 16,293 (100.0) | 13,077 (100.0) | 3,216 (100.0) |
| Antidepressant switched | 918 (5.6) | 758 (5.8) | 160 (5.0) |
| Dose reduced | 634 (3.9) | 608 (4.6) | 26 (0.8) |
| Dose increased | 1,132 (6.9) | 955 (7.3) | 177 (5.5) |
| Dose fluctuated | 413 (2.5) | 386 (3.0) | 27 (0.8) |
| More than one regimen change[f] | 280 (1.7) | 258 (2.0) | 22 (0.7) |
| No changes to drug regimen | 12,916 (79.3) | 10,112 (77.3) | 2,804 (87.2) |
| **Continued a multi-drug[e] regimen throughout pregnancy** | 551 (100.0) | 536 (100.0) | 15 (100.0) |
| Antidepressant added | 97 (17.6) | 93 (17.4) | <5 |
| Antidepressant dropped | 82 (14.9) | 77 (14.4) | 5 (33.3) |
| Products added & dropped | 84 (15.2) | 82 (15.3) | <5 |
| Dose changes | 26 (4.7) | 26 (4.9) | <5 |
| Multiple changes (to dose & product) | 87 (15.8) | 87 (16.2) | <5 |
| No changes to drug regimen | 326 (21.0) | 314 (21.1) | 12 (19.7) |

[a] Those who had at least one prescription for antidepressants in the 3 months prior to pregnancy and during pregnancy
[b] Those who did not have a prescription for antidepressants in the 3 months prior to pregnancy but at least one prescription during pregnancy
[c] Evidence of regimen changes before discontinuation *n*=957 (9.6%)

[d] Those who had an overlapping prescription with the end of pregnancy
[e] Those prescribed at least two, differing antidepressant products >5 days from the end of their current prescription
[f] Those who experienced a switch in product as well as at least one dose change

#### 2.7 Patterns among those with HES

**Table S8** Patterns of prescribing stratified by pre-pregnancy use among those with linked secondary care (HES) data.

| Pattern of prescribing during pregnancy | Total prescribed during pregnancy | 'Prevalent' user[a] | 'Incident' user[b] |
|---|---|---|---|
| **All** | 33,736 | 26,410 | 7,326 |
| Discontinued during pregnancy[c] | 19,044 (56.5) | 14,611 (55.3) | 4,433 (60.5) |
| Continued[d] a single drug regimen throughout pregnancy | 14,078 (41.7) | 11,217 (42.5) | 2,861 (39.1) |
| Continued[d] a multi-drug[e] regimen throughout pregnancy | 614 (1.8) | 582 (2.2) | 32 (0.4) |
| **Continued a single drug regimen throughout pregnancy** | 14,078 (100.0) | 11,217 (100.0) | 2,861 (100.0) |
| Antidepressant switched | 1,247 (8.9) | 1,026 (9.1) | 221 (7.7) |
| Dose reduced | 908 (6.4) | 870 (7.8) | 38 (1.3) |
| Dose increased | 885 (6.3) | 689 (6.1) | 196 (6.9) |
| Dose fluctuated | 942 (6.7) | 881 (7.9) | 61 (2.1) |
| More than one regimen change[f] | 574 (4.1) | 526 (4.7) | 48 (1.7) |
| No changes to drug regimen | 9,522 (67.6) | 7,225 (64.4) | 2,297 (80.3) |
| **Continued a multi-drug[e] regimen throughout pregnancy** | 614 (100.0) | 582 (100.0) | 32 (100.0) |
| Antidepressant added | 614 (100.0) | 582 (100.0) | 32 (100.0) |
| Antidepressant dropped | 106 (17.3) | 99 (17.0) | 7 (21.9) |
| Products added & dropped | 88 (14.3) | 81 (13.9) | 7 (21.9) |
| Dose changes | 121 (19.7) | 113 (19.4) | 8 (25.0) |
| Multiple changes (to dose & product) | 29 (4.7) | 29 (5.0) | <5 |
| No changes to drug regimen | 143 (23.3) | 138 (23.7) | 5 (15.6) |

[a] Those who had at least one prescription for antidepressants in the 3 months prior to pregnancy and during pregnancy
[b] Those who did not have a prescription for antidepressants in the 3 months prior to pregnancy but at least one prescription during pregnancy
[c] Evidence of regimen changes before discontinuation *n*=2,729 (14.3%)
[d] Those who had an overlapping prescription with the end of pregnancy
[e] Those prescribed at least two, differing antidepressant products >5 days from the end of their current prescription
[f] Those who experienced a switch in product as well as at least one dose change

#### 2.8 POST-PREGNANCY PRESCRIBING

**Table S9** Proportion of patients who initiated or resumed antidepressant treatment in the 12 months after pregnancy.

| Previous prescription pattern | Total N | Prescribed after pregnancy n (%) | Not prescribed after pregnancy n (%) |
|---|---|---|---|
| All | 1,033,783 (100.0) | 162,947 (15.8) | 870,836 (84.2) |
| Continued throughout pregnancy | 34,916 (100.0) | 34,916 (100.0) | – |
| Discontinued during pregnancy | 44,228 (100.0) | 23,457 (53.0) | 20,771 (47.0) |
| Discontinued in the 12 months prior to pregnancy | 74,559 (100.0) | 25,532 (34.2) | 49,027 (65.8) |
| No use before or during pregnancy | 880,080 (100.0) | 79,042 (9.0) | 801,038 (91.0) |

**Table S10** Proportion of individuals who were prescribed antidepressants in the 12 months after pregnancy stratified by previous use, among deliveries (postnatal use) and losses (post-pregnancy use)

| Previous prescription pattern | Total N | Prescribed after pregnancy n (%) | Not prescribed after pregnancy n (%) |
|---|---|---|---|
| Deliveries | | | |
| All | 747,311 (100.0) | 113,506 (15.2) | 633,805 (84.8) |
| Continued throughout pregnancy | 18,072 (100.0) | 18,072 (100.0) | – |
| Discontinued during pregnancy | 34,234 (100.0) | 17,642 (51.5) | 16,592 (48.5) |
| Discontinued in the 12 months prior to pregnancy | 51,059 (100.0) | 17,789 (34.8) | 33,270 (65.2) |
| No use before or during pregnancy | 643,946 (100.0) | 60,003 (9.3) | 583,943 (90.7) |
| Losses | | | |
| All | 286,472 (100.0) | 49,441 (17.3) | 237,031 (82.7) |
| Continued throughout pregnancy | 16,844 (100.0) | 16,844 (100.0) | – |
| Discontinued during pregnancy | 9,994 (100.0) | 5,815 (58.2) | 4,179 (41.8) |
| Discontinued in the 12 months prior to pregnancy | 23,500 (100.0) | 7,743 (32.9) | 15,757 (67.1) |
| No use before or during pregnancy | 236,134 (100.0) | 19,039 (8.1) | 217,095 (91.9) |

**Table S11** Proportion of individuals who initiated or resumed antidepressant treatment in the 12 months after pregnancy among those that had at least 12 months follow-up post-pregnancy.

| **Previous prescription pattern** | **Total N** | **Prescribed after pregnancy n (%)** | **Not prescribed after pregnancy n (%)** |
|---|---|---|---|
| All | 921,521 (100.0) | 151,454 (16.4) | 770,067 (83.6) |
| Continued throughout pregnancy | 31,105 (100.0) | 31,105 (100.0) | – |
| Discontinued during pregnancy | 38,992 (100.0) | 21,742 (55.8) | 17,250 (44.2) |
| Discontinued in the 12 months prior to pregnancy | 65,816 (100.0) | 23,849 (36.2) | 41,967 (63.8) |
| No use before or during pregnancy | 785,608 (100.0) | 74,758 (9.5) | 710,850 (90.5) |

#### 2.9 INDICATIONS

**Table S12** Incident depression and anxiety at different times in relation to pregnancy.

| Indication | Total N | Prescribed during pregnancy n (%) | Not prescribed during pregnancy n (%) |
|---|---|---|---|
| | 1,033,783 (100.0) | 79,144 (100.0) | 954,639 (100.0) |
| **Depression ever before the end of pregnancy** | | | |
| Depression started more than 12 months before pregnancy | 223,775 (21.6) | 49,447 (62.5) | 174,328 (18.3) |
| Depression started 12 months before pregnancy | 31,339 (3.0) | 10,482 (13.2) | 20,857 (2.2) |
| Depression started during pregnancy | 7,949 (0.8) | 3,866 (4.9) | 4,083 (0.4) |
| **Anxiety ever before the end of pregnancy** | | | |
| Anxiety started more than 12 months before pregnancy | 136,469 (13.2) | 29,941 (37.8) | 106,528 (11.2) |
| Anxiety started 12 months before pregnancy | 19,661 (1.9) | 5,937 (7.5) | 13,724 (1.4) |
| Anxiety started during pregnancy | 7,985 (0.8) | 2,388 (3.0) | 5,597 (0.6) |

**Table S13** Prevalence of other indications among those prescribed to antidepressants during pregnancy with depression and/or anxiety.

| | Total prescribed n (%) | Depression n (%) | Anxiety n (%) | Depression and anxiety n (%) | Neither depression nor anxiety n (%) |
|---|---|---|---|---|---|
| **Total** | 79,144 (100.0) | 32,204 (100.0) | 6,675 (100.0) | 31,591 (100.0) | 8,674 (100.0) |
| Depression | 63,795 (80.6) | 32,204 (100.0) | - | 31,591 (100.0) | - |
| Anxiety | 38,266 (48.3) | - | 6,675 (100.0) | 31,591 (100.0) | - |
| Other mood disorders* | 51 (0.1) | 22 (0.1) | <5 | 27 (0.1) | <5 |
| Eating disorders | 4,142 (5.2) | 1,468 (4.6) | 234 (3.5) | 2,177 (6.9) | 263 (3.0) |
| Pain | 7,263 (9.2) | 2,542 (7.9) | 482 (7.2) | 3,544 (11.2) | 695 (8.0) |
| Diabetic neuropathy | 28 (0.0) | 16 (0.0) | <5 | 7 (0.0) | <5 |
| Stress incontinence | 2,213 (2.8) | 834 (2.6) | 124 (1.9) | 1,145 (3.6) | 110 (1.3) |
| Migraine prophylaxis | 638 (0.8) | 215 (0.7) | 46 (0.7) | 308 (1.0) | 69 (0.8) |
| Tension-type headache | 88 (0.1) | 34 (0.1) | <5 | 43 (0.1) | 7 (0.1) |
| No evidence of an antidepressant indication | 7,578 (9.6) | - | - | - | 7,578 (87.4) |

#### 2.10 Missing data

**Table S14** Likelihood of having missing data in ethnicity, BMI, smoking status, alcohol use status having discontinued.

| Variable | Discontinuers with missing | Continuers with missing | Likelihood of missing data [aOR[a] (95% CI)] |
|---|---|---|---|
| Maternal ethnicity | 14,490/44,228 (32.76) | 11,074/34,916 (31.72) | 1.03 (1.00-1.06) |
| Maternal BMI around the start of pregnancy | 3,868/44,228 (8.75) | 2,668/34,916 (7.64) | 1.18 (1.12-1.25) |
| Smoking status around the start of pregnancy | 1,431/44,228 (3.24) | 812/34,916 (2.33) | 1.11 (1.01-1.21) |
| Alcohol consumption around the start of pregnancy | 10,110/44,228 (22.86) | 7,181/34,916 (20.57) | 1.14 (1.10-1.18) |

[a] adjusted for year of pregnancy start
